# STABILITY ANALYSIS OF DISEASE FREE EQUILIBRIUM STATE (*E*_0_) FOR AN ENDEMIC DETERMINISTIC MODEL FOR HIV/AIDS

**DOI:** 10.1101/2020.03.31.20048918

**Authors:** A. O. Victor, H. K. Oduwole

**Affiliations:** Department of Mathematics, Nasarawa State University, Keffi, Nigeria

**Keywords:** Stability, Asymptotical, SEURUS, Disease-free equilibrium

## Abstract

This paper focused on determining the asymptotical stability of the new model which establishes that a disease-free equilibrium state exists and is locally asymptotically stable when the basic reproduction number 0 ≤ *R*_0_ < 1 and the following threshold conditions (0 ≤ *R*_1_ < 1, 0 ≤ *R*_2_ < 1, 0 ≤ *R*_3_ < 1, 0 ≤ *R*_4_ < 1 and 0 ≤ *R*_5_ < 1, 0 ≤ *R*_6_ < 1, 0 ≤ *R*_7_ < 1, 0 ≤ *R*_8_ < 1) are satisfied. Results from the model analysis shows that the proportion of infected juvenile and adult sub-population in the presence of High Active Anteritroviral Therapy (HAART) drastically reduce to a zero (*R*_0_ = 0) as compared to the infected age-structured population when treatment rate is low and the net transmission rate is near zero.

## 1. INTRODUCTION

In this paper, the stability theory is investigated and it addresses the stability of solutions of the ordinary differential equations describing the disease free equilibrium state of a deterministic endemic model for the control of the spread of endemic disease. Solutions to systems of differential equations which model disease transmission are of particular use and importance to epidemiologists interested in studying effective means to slow and prevent spread of the disease (Garnett, 2002).

In dynamic systems, an orbit is called Lyapunov stable if the forward orbit of any point is in a small enough neighborhood or it stays in a small neighborhood. Various criteria has been developed to prove stability or instability or an orbit but under favorable circumstances, the question may be reduced to a well-studied problem involving eigenvalues of matrices. A more general method involves Lyapunov function and in practice, any one of a number of different stability criteria is applied to this study.

Many parts of the qualitative theory of differential equations and dynamical systems deal with asymptotic properties of solutions and the trajectories. Hence, the simplest type of behavior is exhibited by equilibrium points, or fixed points, and by periodic orbits. In this paper we model the age-structured population deterministic model with the various compartmental sub-population from the susceptible group, to the exposed class and from the exposed class to the infectious class and then to the removed (recovered) class and the undetectable class who are actively using the Highly Active Antiretroviral Therapy (HAART) which is a combination of more drug types as an advance therapy from Antiretroviral therapy (ART). The investigation of the deterministic endemic SEIRUS (Susceptible – Exposed – Infectious – Recovered – Undetectable=Untransmitable – Susceptible) model is a novel method in the scientific and empirical approach in controlling the spread of diseases in an endemic situation which is a build up from Mckendrick and Kermack (1927).

## 2. THE MODEL VARIABLES AND PARAMETERS

The model variables and parameters for the investigation of the stability analysis of the equilibrium state for the new deterministic endemic model which is a motivation from Oduwole and Kimbir (2018) is given by;

### 2.1 Variable and Parameters

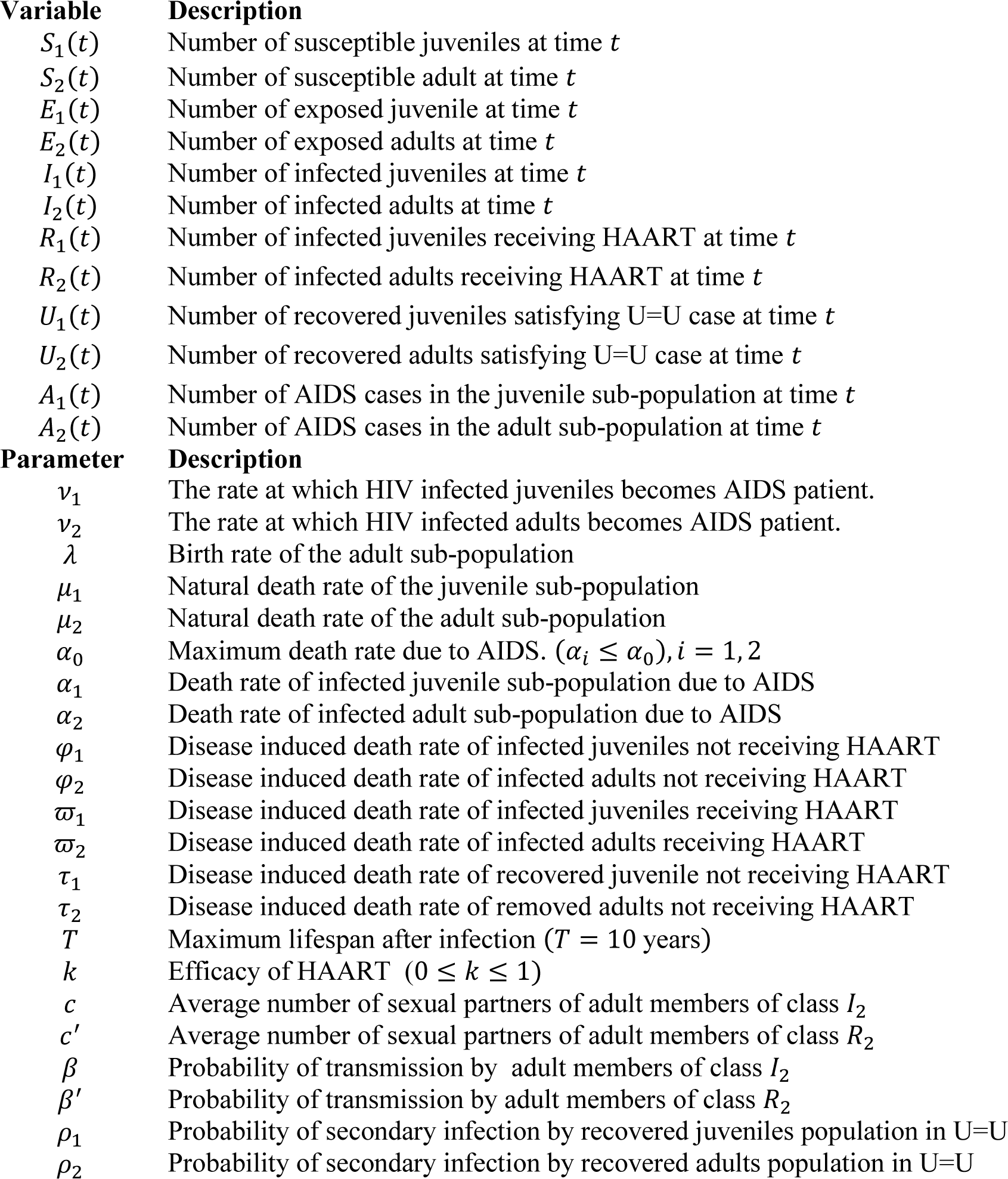
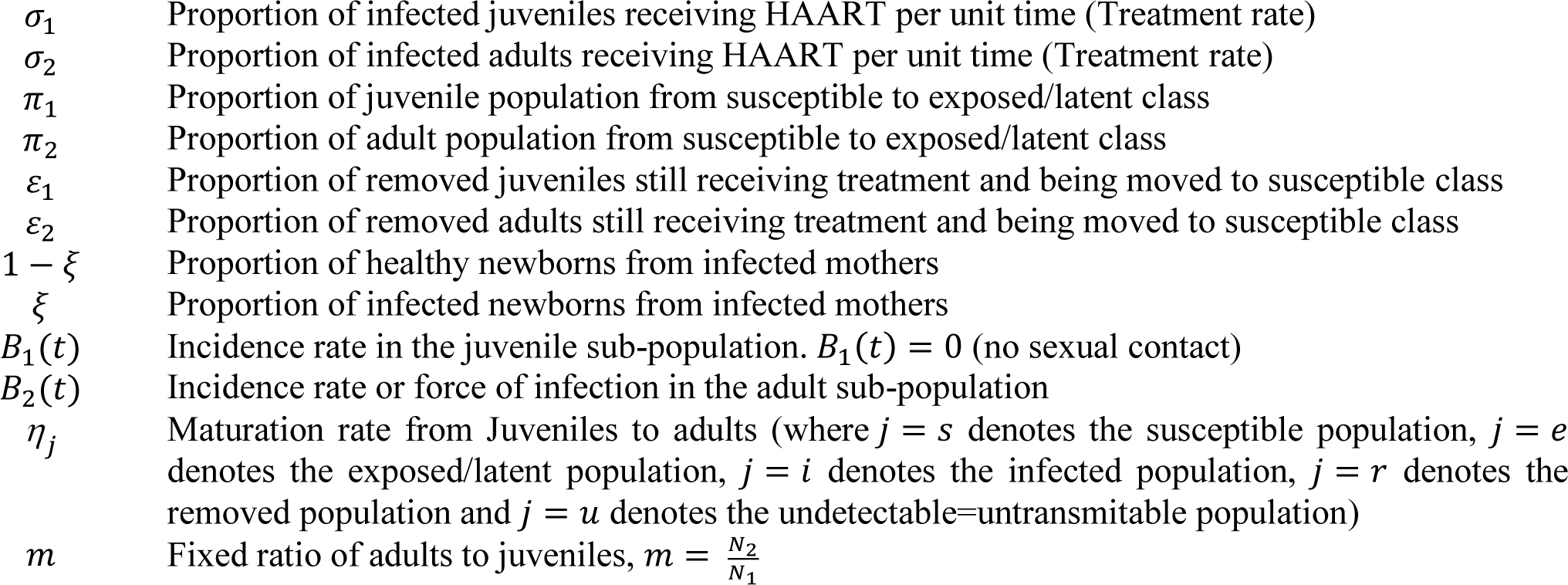

### 2.2 MODEL ASSUMPTIONS

The following assumptions would help in the derivation of the model:

1. There is no emigration from the total population and there is no immigration into the population. A negligible proportion of individuals move in and out of the population at a given time.
2. Maturation (or maturity) is interpreted as growth from Juvenile stage into adult.
3. The susceptible population are first exposed to a latent class where they can infected or not.
4. Some infected individuals move to the removed class when counseled and are placed under highly active antiretroviral therapy (HAART).
5. Newborns are not of the same class as their progenitor. A fraction (1 − *ξ*) of newborns from infected mothers are healthy, while the remaining fractions *ξ* are born with the virus.
6. Only an adult can reproduce.
7. The rate of progression from HIV to AIDS is different for both juvenile and adult sub-populations.
8. The AIDS cases have full-blown symptoms and are therefore not sexually active.
9. The recruitment into the *S*-class is only through birth for the juvenile sub-population and through maturation for the adult sub-population.
10. The recruitment from the *S*-class into the *E*-class is through birth for newborns and through heterosexual activities for adults. This is done at a rate *π*_1_ and *π*_2_ for the juvenile and adult sub-population respectively.
11. The recruitment into the *R*-class from the *I*-class depends on the effectiveness of public campaign and counselling. This is done at a rate *σ*1 and *σ*2 for the juvenile and adult sub-populations, respectively.
12. The recruitment into the *U*-class from the *R*-class depends on the effectiveness of the HAART and the change in social behavior of the recovered population. This is done at a rate *ρ*_1_ and *ρ*_2_ for the recovered juvenile and adult sub-population respectively.
13. The recruitment into the *S*-class over again from the *U*-class depends on how long the population in the *U*-class remain in the class while actively receiving treatment. This stage it is assumed that the compartment is filled with fully removed population whose viral load is less than 1% and have 0% chance of secondary infection. This is done at a rate *ε*_1_ and *ε*_2_ for juvenile and adult sub-population respectively.
14. There is a chance of infection by the juvenile and adult population in the U=U class at *ρ*_1_ and *ρ*_2_ probability if the administration of HAART is discontinued at any given time.
15. Death is implicit in the model and it occurs in all classes at constant rate *μ*_*i*_, where *i* = 1,2 represents the juvenile and adult sub-population respectively. However, there is an additional death rate in the *I* and *R* classes due to infection for both juvenile and adult sub-population denoted by *φ*_*i*_ and *ϖ*_*i*_ respectively, where *i* = 1, 2 represents the juvenile and adult sub-population respectively.
16. There is a maximum period of time, *T* after infection, which a member in class *I* who progresses to AIDS dies. The death rate in the *R*-class is given by *ϖ*_*i*_ = *φ*_*i*_*e*^−*kT*^, where *φ*_*i*_ is the death rate due to the infection, and *k* is the efficacy of the antiretroviral drugs. The higher the value of *k*, the smaller the value of *ϖ*_*i*_ and vice versa. Clearly *ϖ*_*i*_ ≤ *φ*_*i*_ and *ϖ*_*i*_ = *φ*_*i*_ when *k* = 0 (i.e. no HAART). Also *φ*_*i*_ ≤ *α*_0_ (the maximum death rate due to AIDS).
17. There is a fixed ratio of adults to juveniles given by 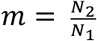. This assumption allow for asuitable control of the population at a given time.

### 2.3 MODEL DESCRIPTION

The susceptible-exposed-infected-removed-undetectable=untransmissible-susceptible (SEIRUS) model that considers an open age-structured population of juvenile and heterosexual individuals is formulated based on the McKendrick-von-Foerster type two-age-structured SIR model as used by Oduwole and Kimbir (2018) in studying the effect of antiretroviral therapy. The population is sub-divided based on demographic structure and epidemiological structure. Under a demographic structure, the population is divided into classes, the juvenile class (0 – 14 years) and the adult class (15 years and above), while under the epidemiological structure of this study is divided into six classes, namely; susceptible (*S*), exposed (E), infected (*I*), removed (*R*), Undetectable=Untransmitable (U=U) and those infected progressing to AIDS **(*A*)**. A susceptible is an individual that is yet to be infected, but is open to infection as he or she interacts with members of the *I*-class. An infected individual is one who has contracted HIV and is at some stage of infection. A removed individual is one that is confirmed to be HIV positive, counseled, and is receiving treatment via highly active antiretroviral therapy (HAART) (Murray, *et. al*., 1998). A member of the Undetectable=Untransmitable class is one that has been removed and has been actively receiving treatment through HAART and has been satisfied by the UN-MDG 6’s standard to be in the U=U class. A member of the ***A***-class is an individual who is HIV positive, and has progressed to full blown AIDS (CDC, 2018).

The following algorithm describes the dynamic of SEIRUS framework, and will be useful in the formulation of model equations.

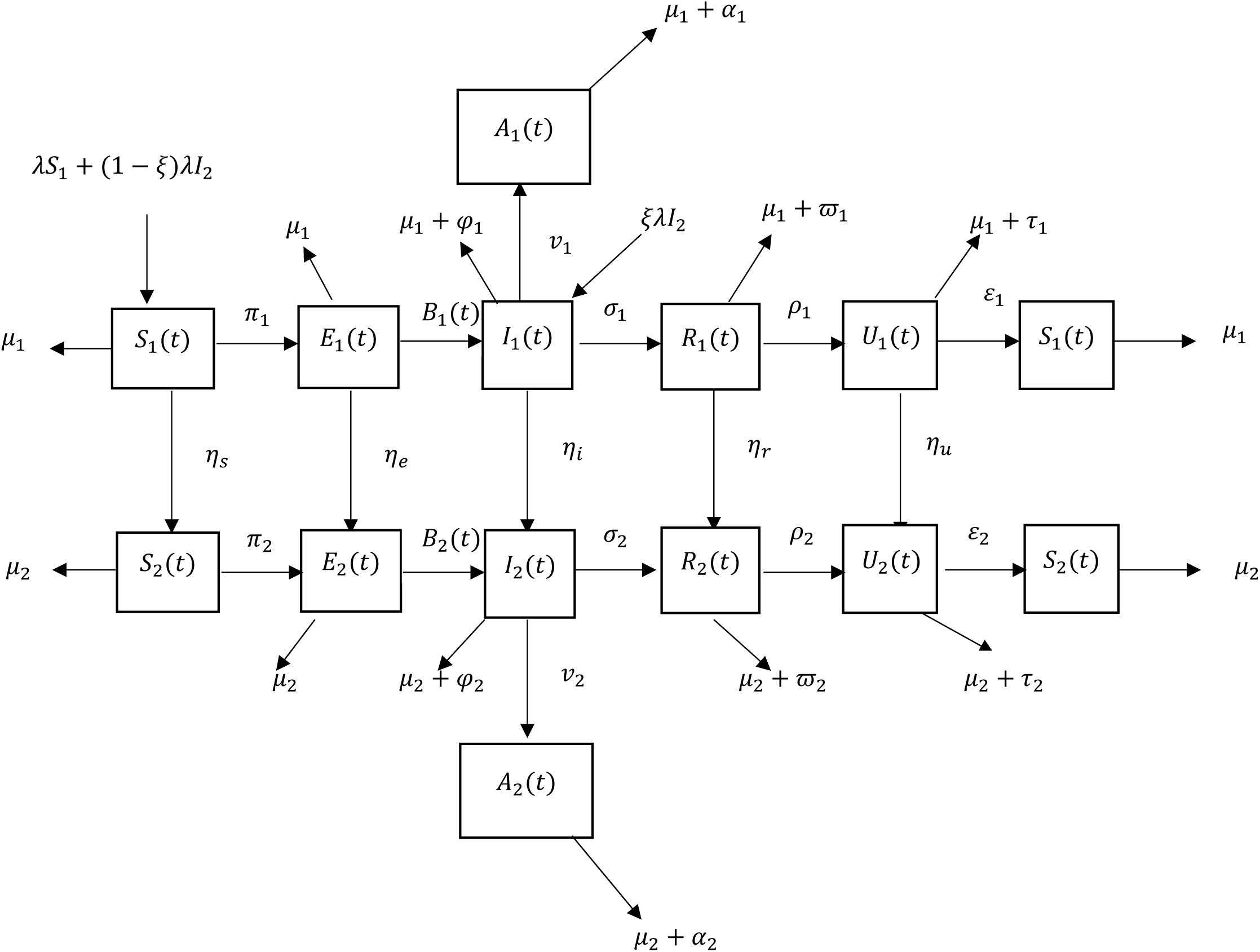

## 3. THE MODEL EQUATIONS

From the assumptions and the flow diagram above, the following model equations are derived.

**For the Juvenile sub-populations:**

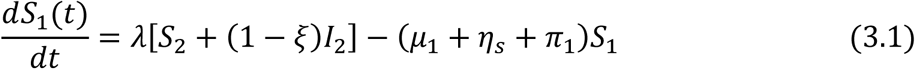

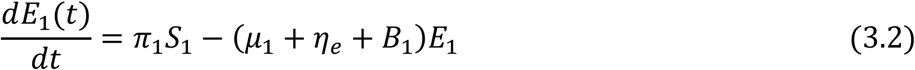

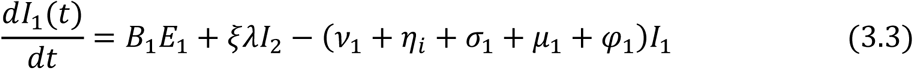

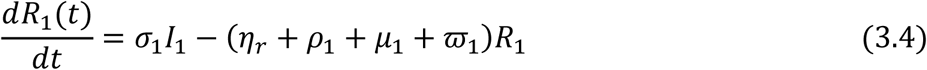

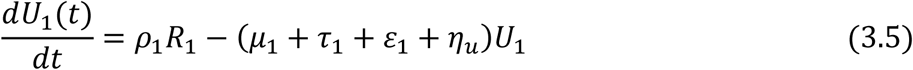

**For the Adult sub-populations:**

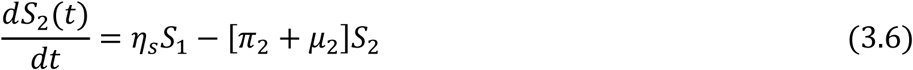

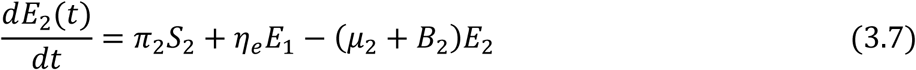

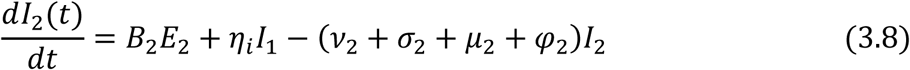

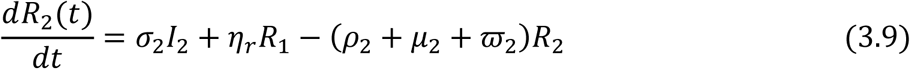

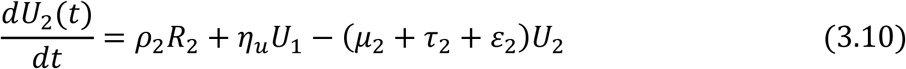

**For those progressing to AIDS:**

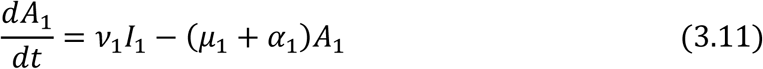

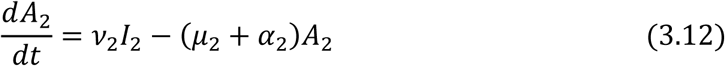

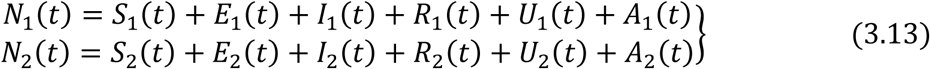

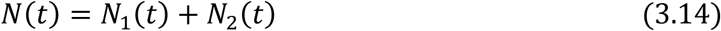

The incidence rate or force of infection at time *t* denoted by *B*_2_(*t*) in the adult population is given as

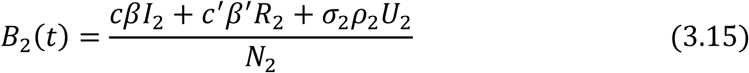

### 3.1 MODEL EQUATIONS IN PROPORTIONS

To simplify the model, it is reasonable to assume that infected juvenile and adult who progress to full blown AIDS are isolated and sexually inactive; hence they are not capable of producing children (vertical transmission) and they do not contribute to viral transmission horizontally (from adult to adult).

To achieve this, we normalize the model by transforming the model equations into proportions and eliminate the AIDS class ***A*(***t*), which invariably reduces the number of model equations from twelve to ten. The derive model equations in proportion of infected juveniles and adults define prevalence of infection, which has biological meaning.

The model equations are transformed into proportions as follows;

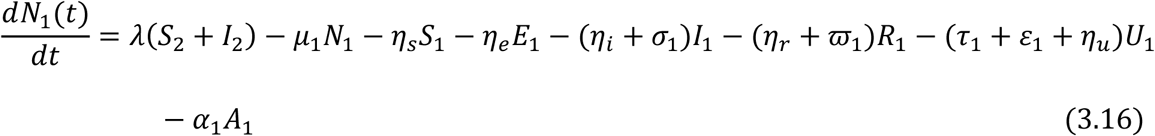

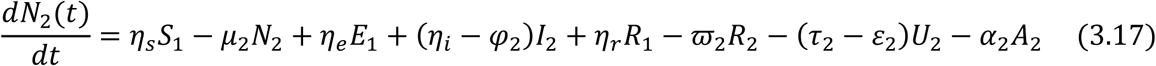

Let

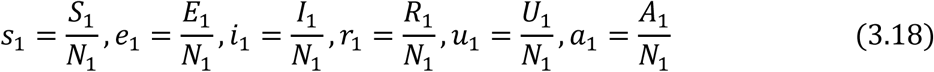

similarly,

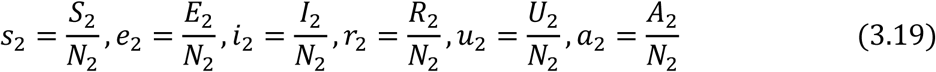

and

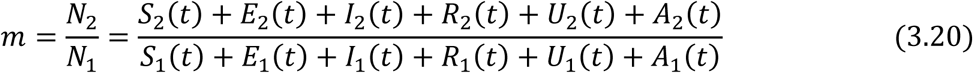

Then the normalized system can be calculated as follows,

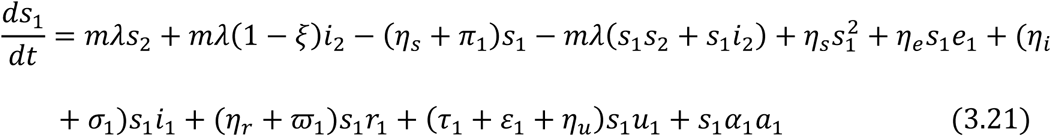

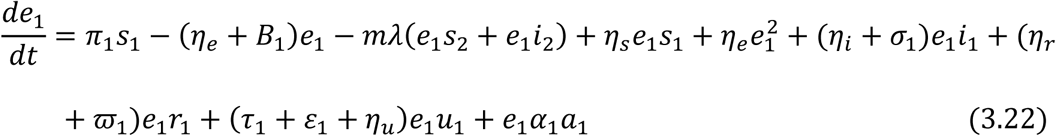

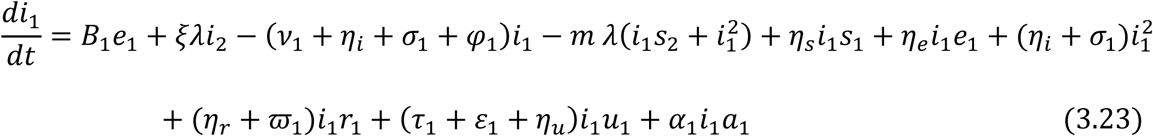

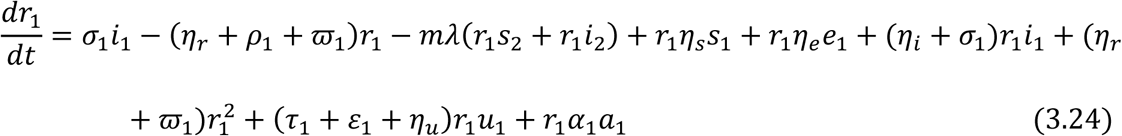

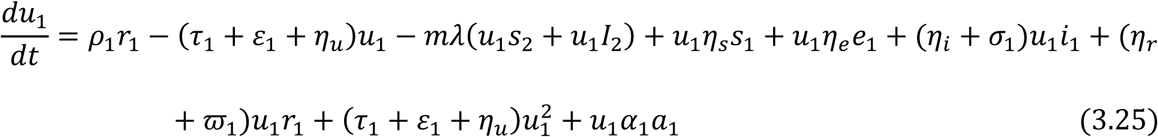

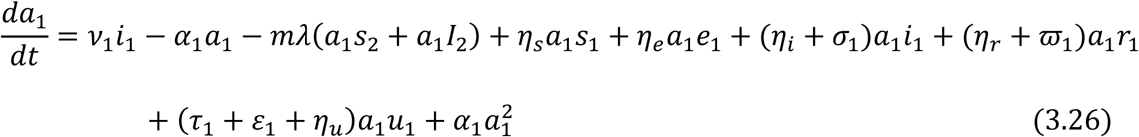

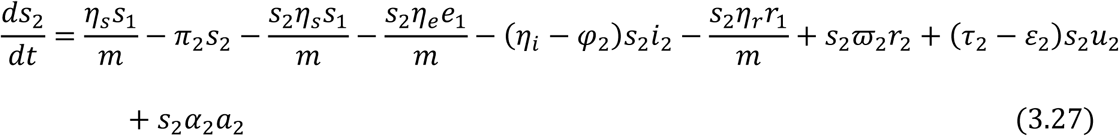

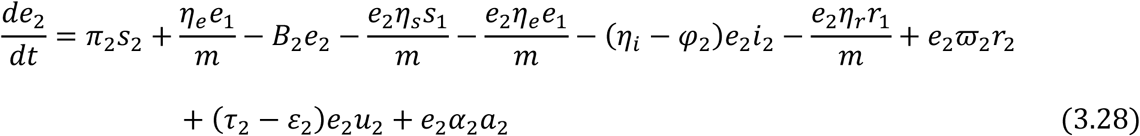

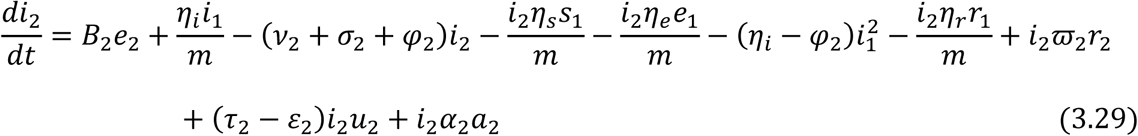

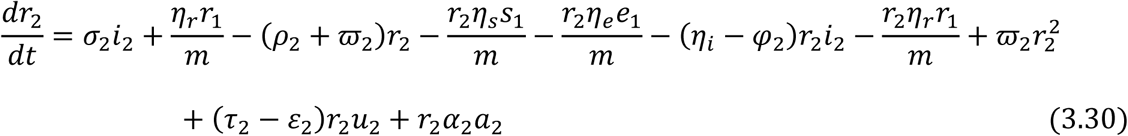

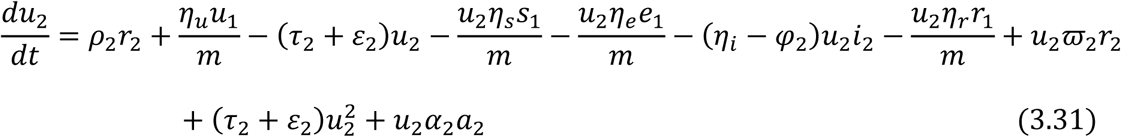

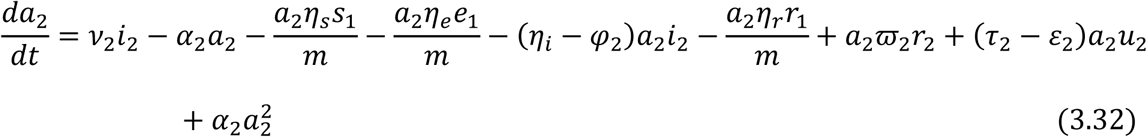

From equation (3.18) and (3.19), we have

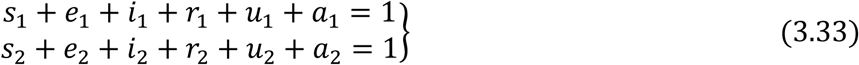

Therefore substituting *a*_1_ = 1 − (*s*_1_ + *e*_1_ + *i*_1_ + *r*_1_ + *u*_1_) and *a*_2_ = 1 − (*s*_2_ + *e*_2_ + *i*_2_ + *r*_2_ + *u*_2_) in equations (3.21) – (3.32) respectively gives the following governing equations of the model below:

For the Juvenile sub-population:

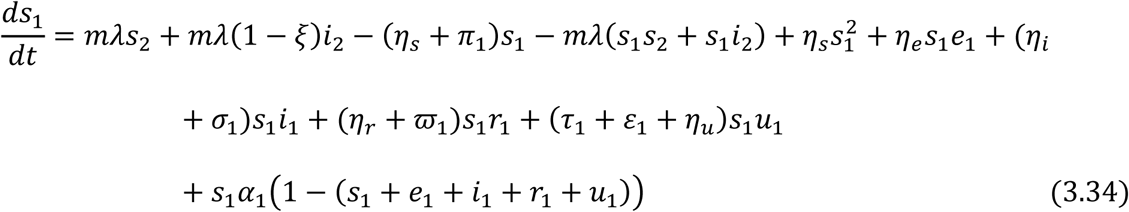

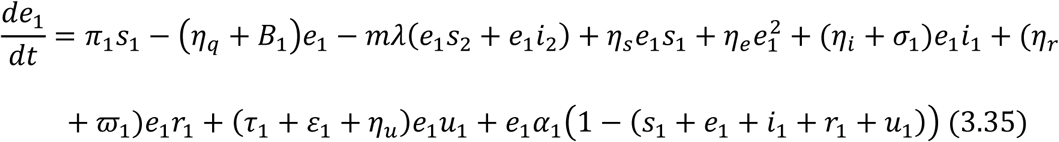

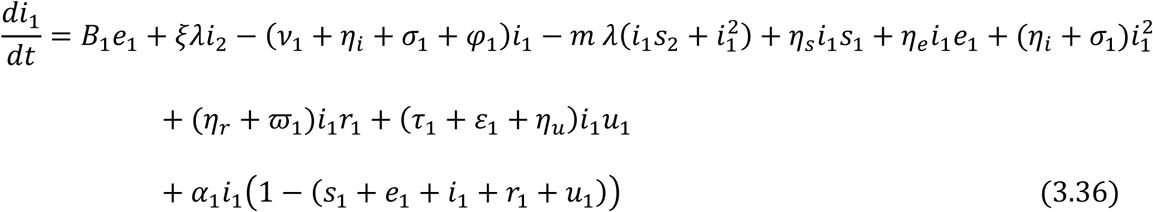

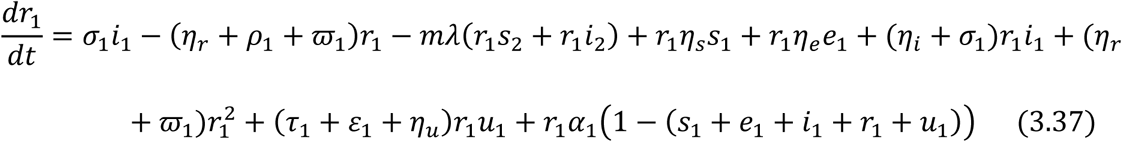

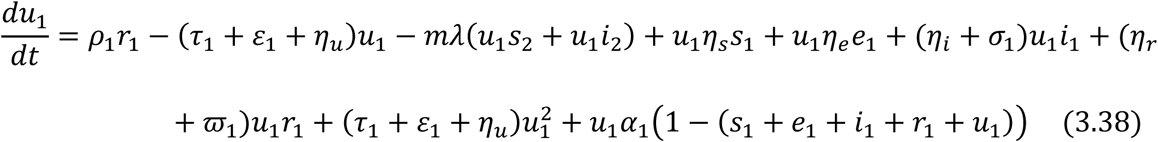

For the Adult sub-population:

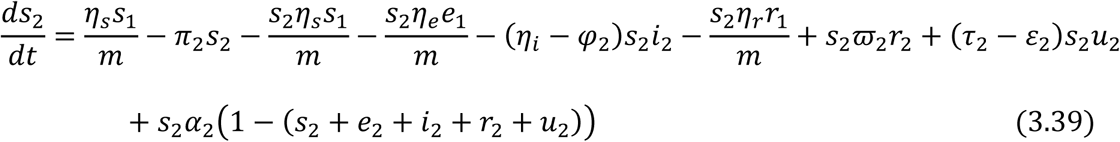

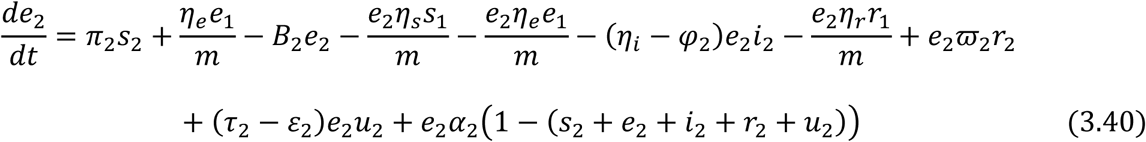

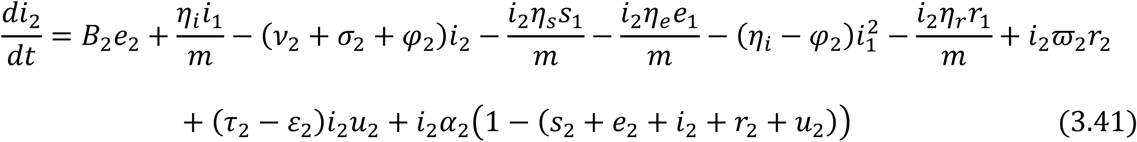

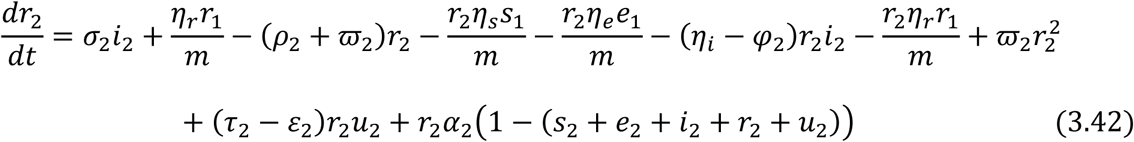

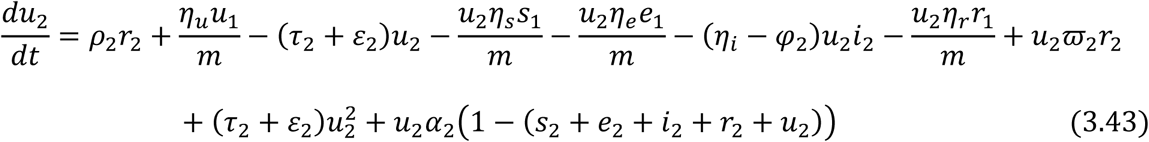

Equations (3.34) to (3.43) are the model equations in proportions, which define prevalence of infection.

### 3.2 EXISTENCE AND UNIQUENESS OF DISEASE FREE EQUILIBRIUM STATE (*E*_0_) OF THE SEIRUS MODEL

The disease-free equilibrium (DFE) state of the endemic SEIRUS model is obtained by setting the left hand sides of equations (3.34) – (3.43) to zero and setting the disease components *e*_1_ = 0, *e*_2_ = 0, *i*_1_ = 0, *i*_2_ = 0, *r*_1_ = 0, *r*_2_ = 0 and *u*_1_ = 0, *u*_2_ = 0 leading to equations (3.44) – (3.45) below

For the Juvenile sub-population:

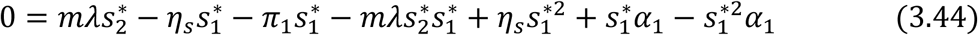

For the Adult sub-population:

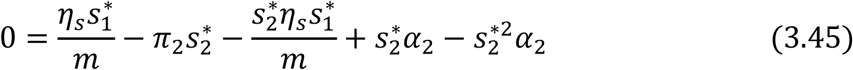

Factorizing 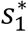 From Equation (3.45) and substituting into (3.44) gives;

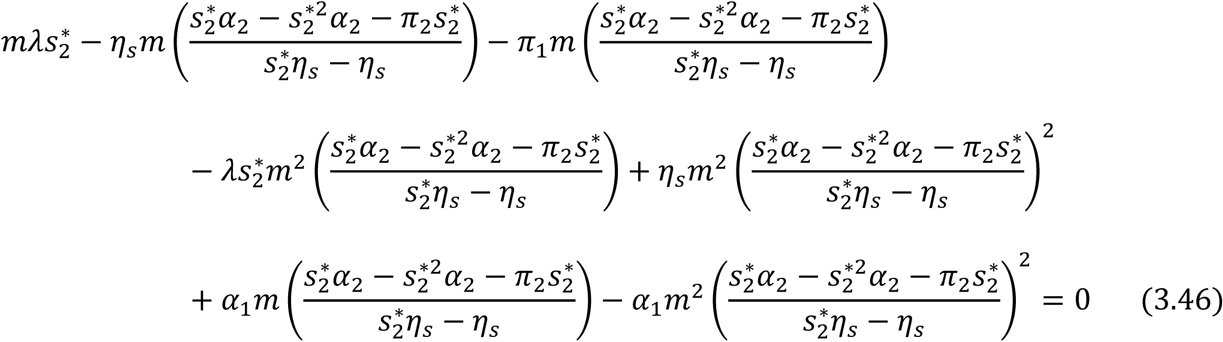

Multiplying through by 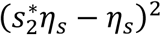, gives

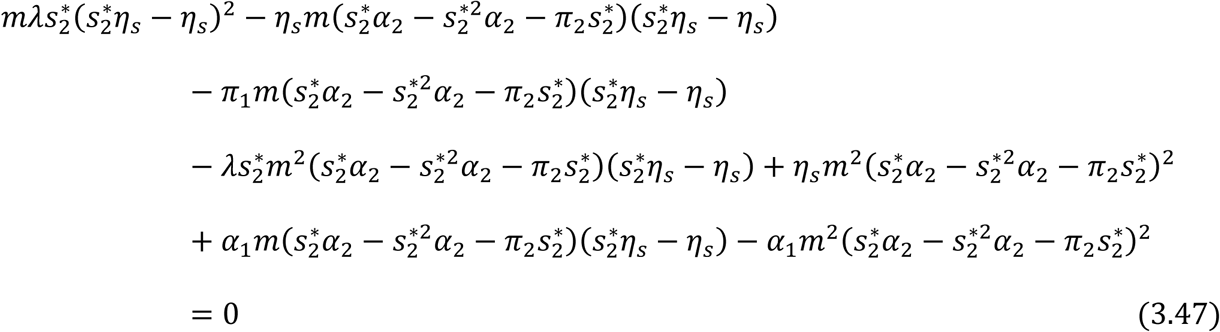

Simplifying further and collecting like terms in 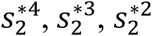 and 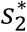 gives,

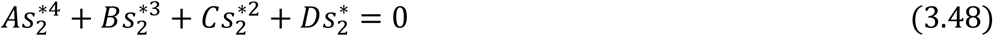

where

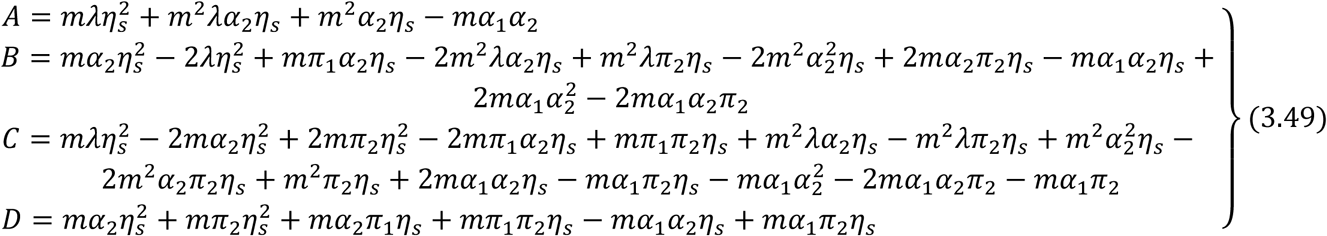

Factorizing 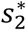 in (3.48) gives;

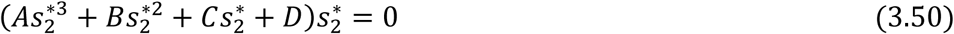

Therefore, the solution for the simultaneous equations (3.50) with (3.49) is given by

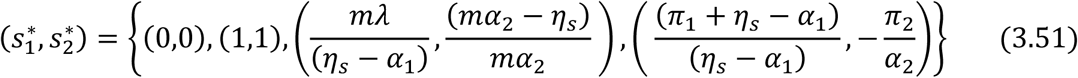

Ignoring the native values of 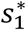 and 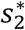 and other stringent conditions, there exist a unique trivial and disease-free equilibrium states at 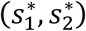 given by (0,0) and (1,1) respectively.

The solution (3.51) satisfies equations (3.48) and (3.47) identically.

## 4. STABILITY ANALYSIS OF DISEASE FREE EQUILIBRIUM STATE (*E*_0_)

To study the behavior of the system (3.34) – (3.43) around the disease-free equilibrium state *E*_0_ = [1,0,0,0,0,1,0,0,0,0], we resort to the linearized stability approach.

Let *f*_*ji*_, *i* = 1,2,3,4,5 be used to linearize the proportional governing equations for the juvenile sub-population and *f*_*ai*_, *i* = 1,2,3,4,5 be used to linearized the proportional governing equations for the adult sub-population.

For the Juvenile sub-population:

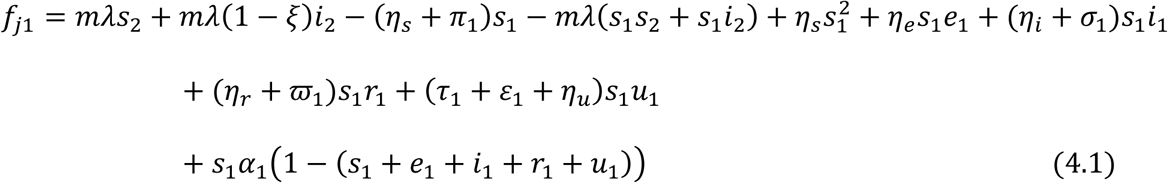

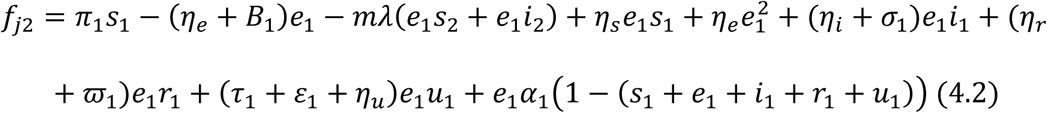

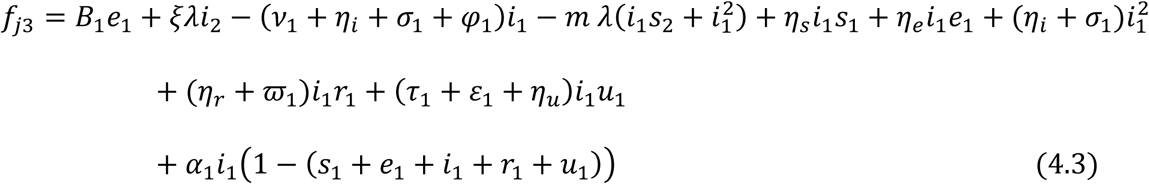

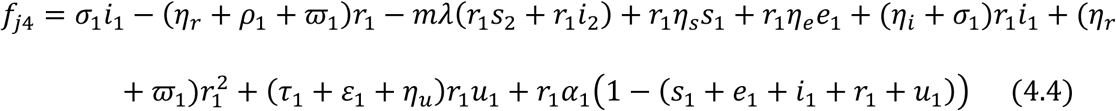

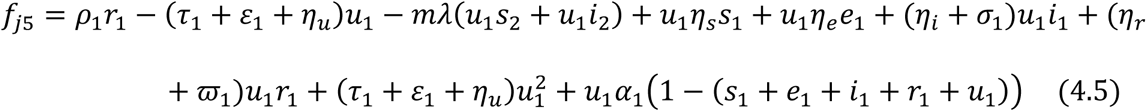

For the Adult sub-population:

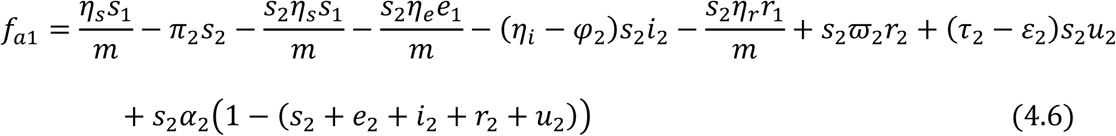

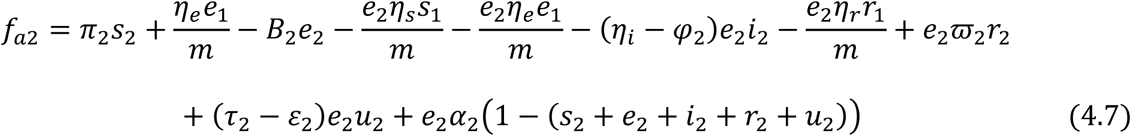

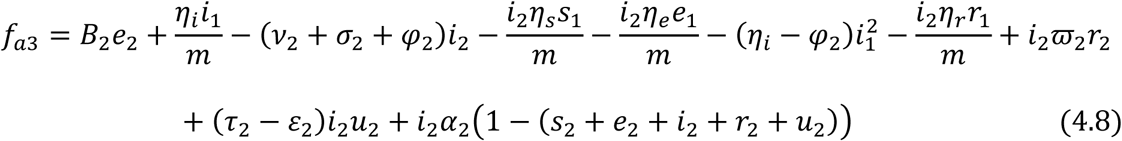

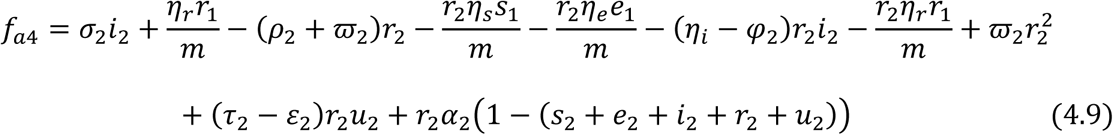

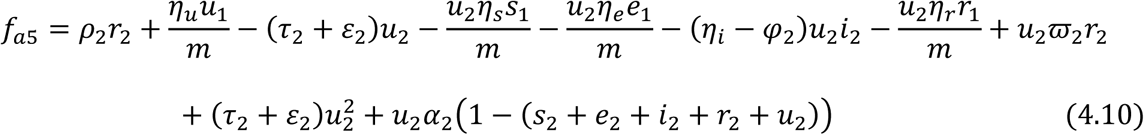

Then

For the Juvenile sub-population:

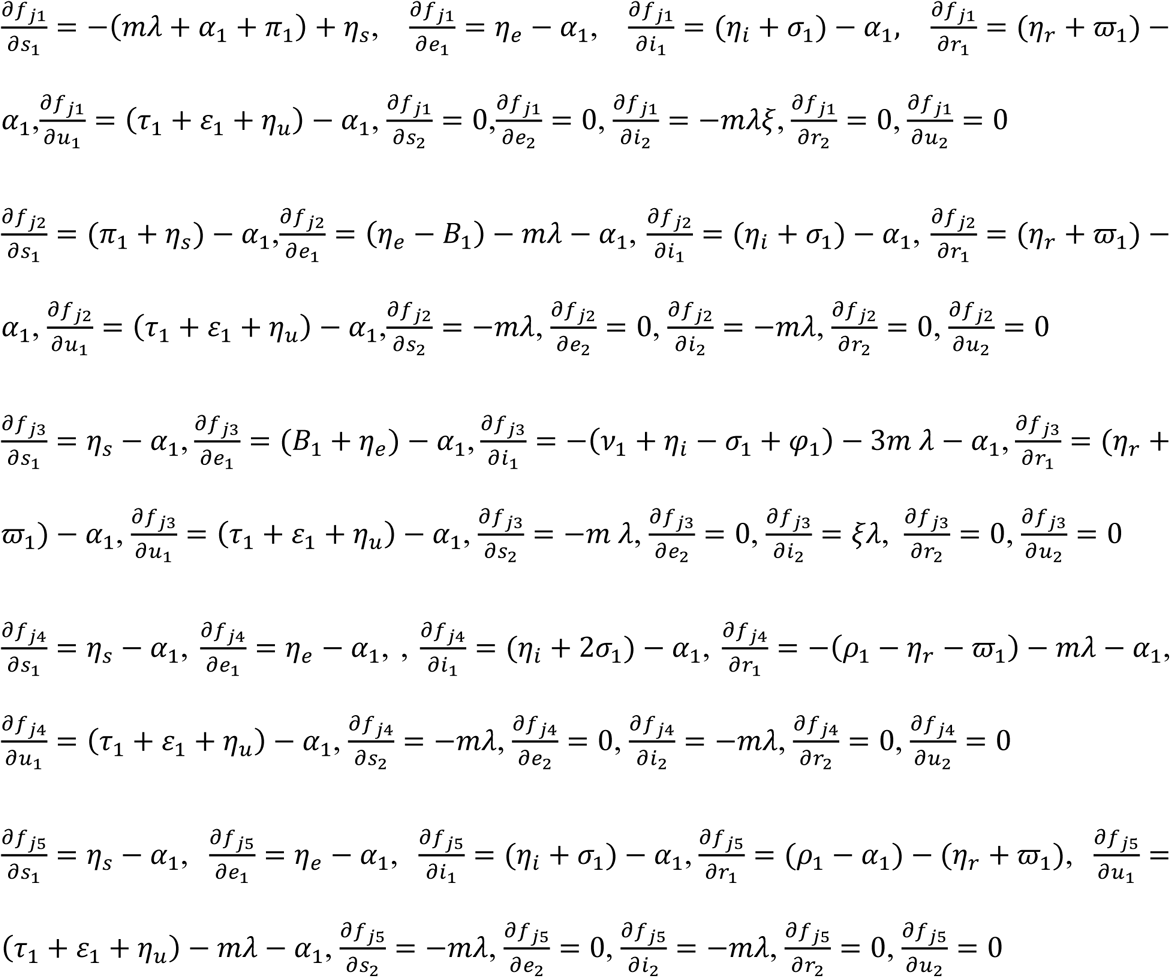

For the Adult sub-population:

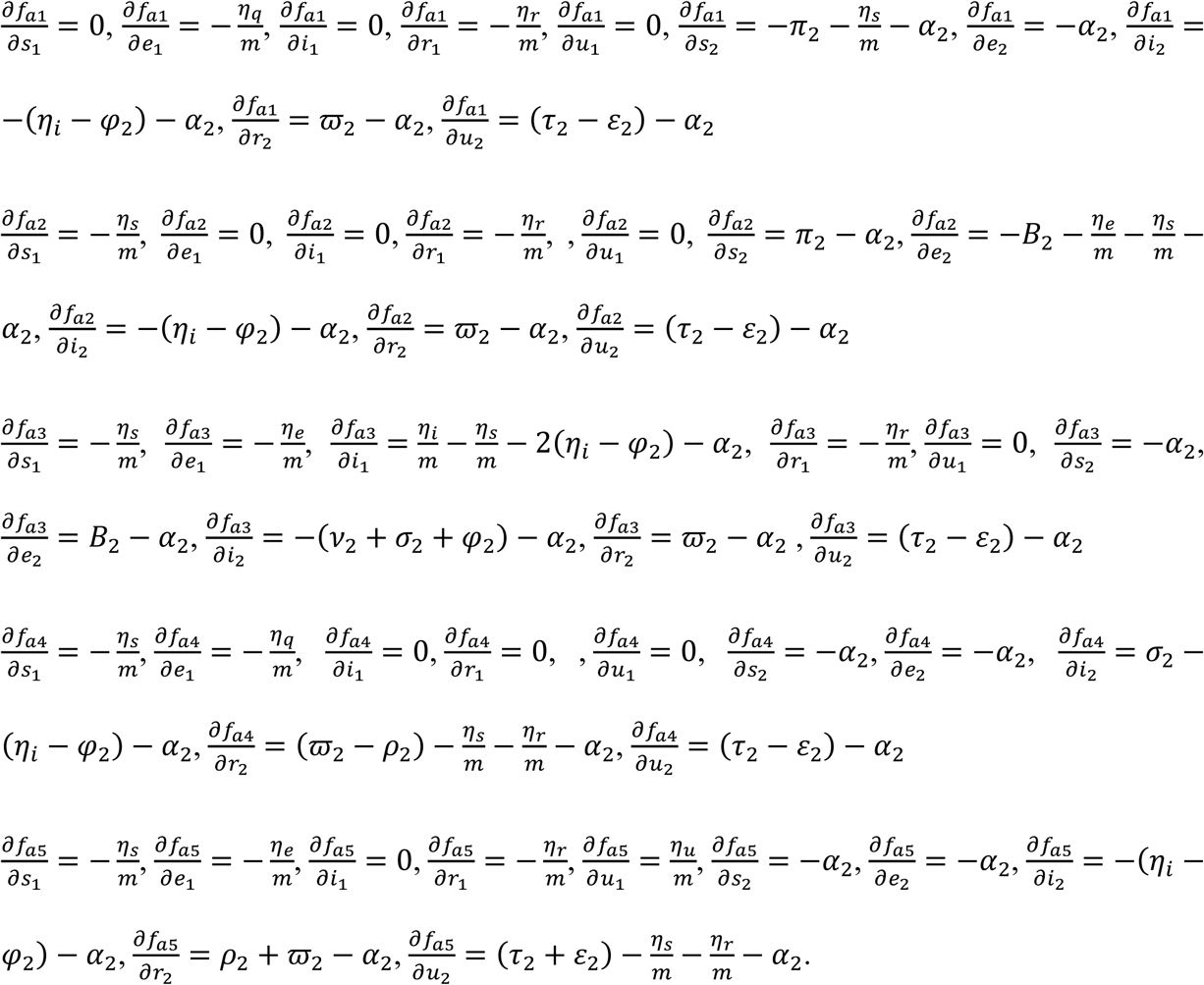

The Jacobian 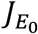 is given by

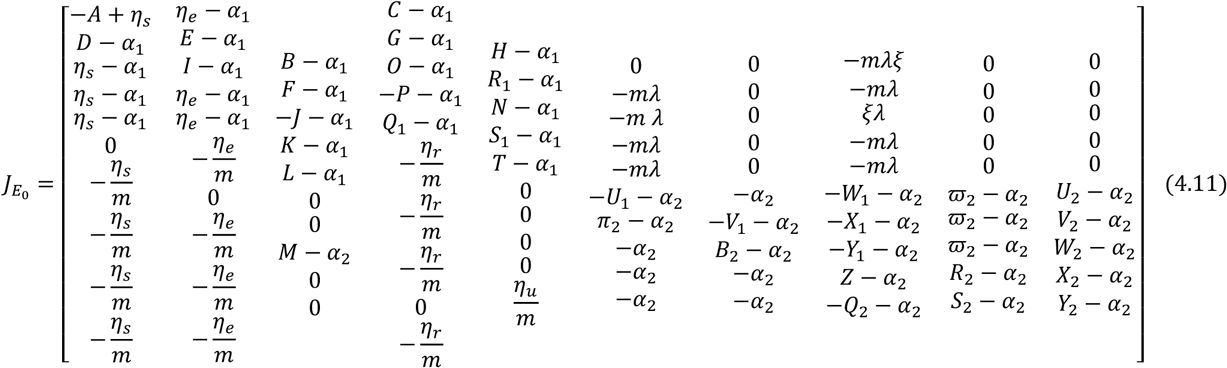

where

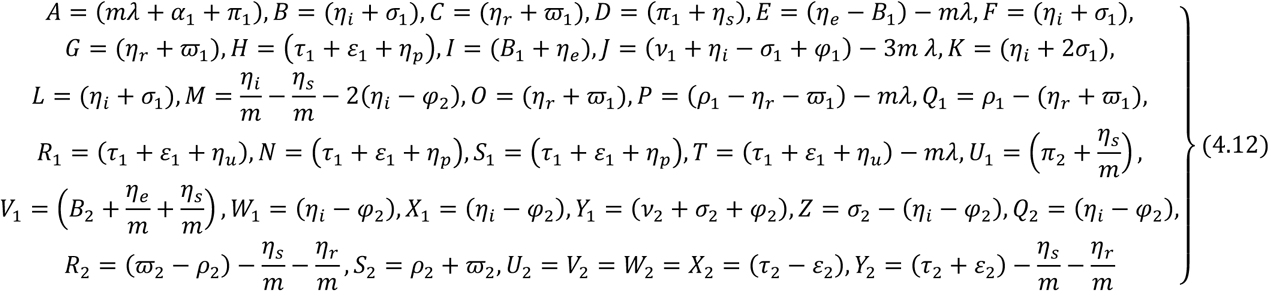

According to Silvester (1999), the determinant of the Jacobian Matrix 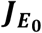 is given by the recursive definition for a 10 × 10 matrix defined as;

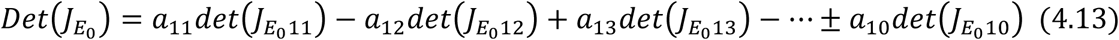

Which is used to resolve the 10×10 matrix in (4.14) and hence we get;

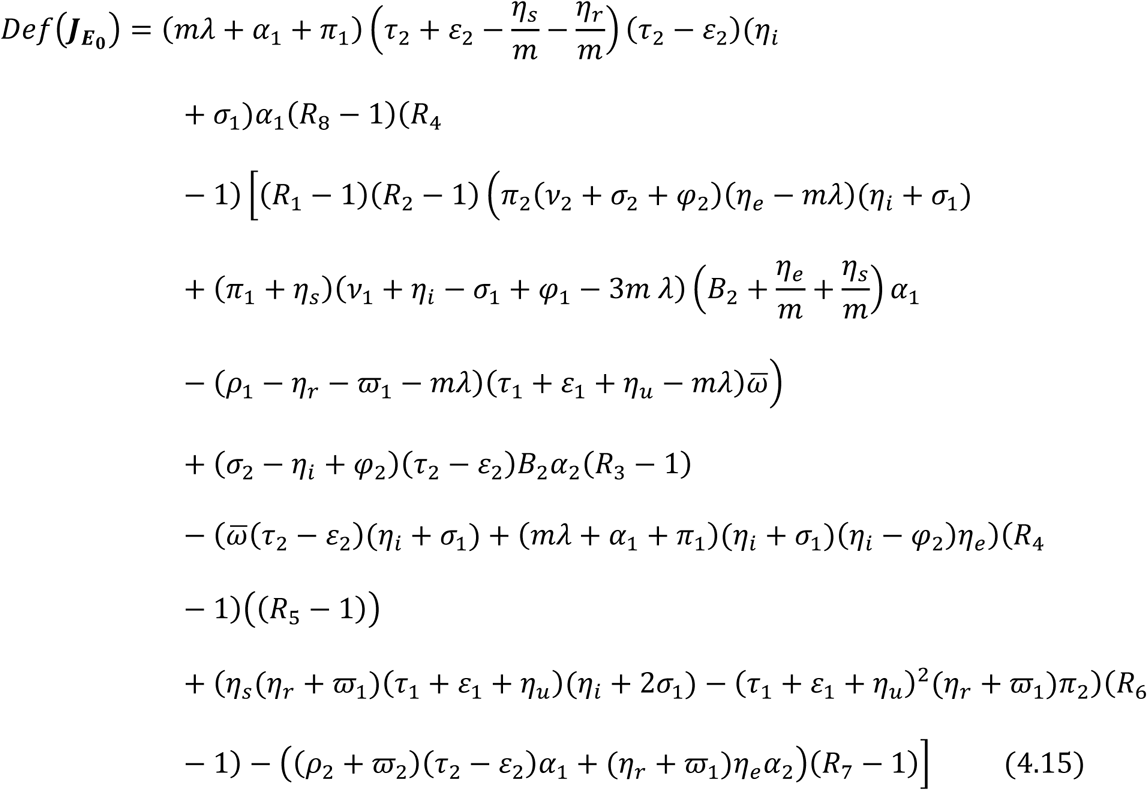

where

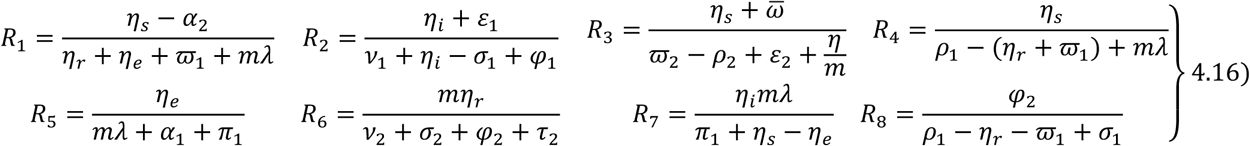

Similarly, the Trace of the Jacobian Matrix 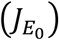 is given by

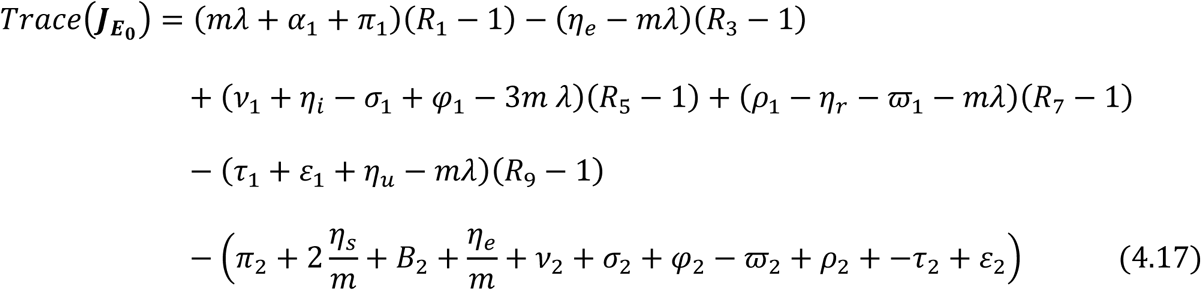

where

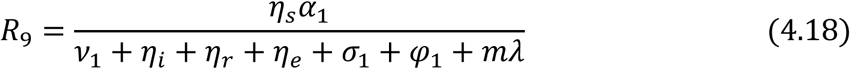

**Theorem 1:** The disease free equilibrium state *E*_0_ = [1,0,0,0,0,1,0,0,0,0] of the model (3.34) – (3.43) is locally asymptotically stable if *R*_0_ < 1 and the following conditions hold

(i). *R*_1_ < 1 (ii). *R*_2_ < 1 (iii). *R*_3_ < 1 (iv). *R*_4_ < 1 (v). *R*_5_ < 1 (vi). *R*_6_ < 1 (vii). *R*_7_ < 1 (viii). *R*_8_ < 1 (ix). *R*_9_ < 1 otherwise *E*_0_ is unstable.

### Proof

The Jacobian matrix of the system (3.34) – (3.48) is given by

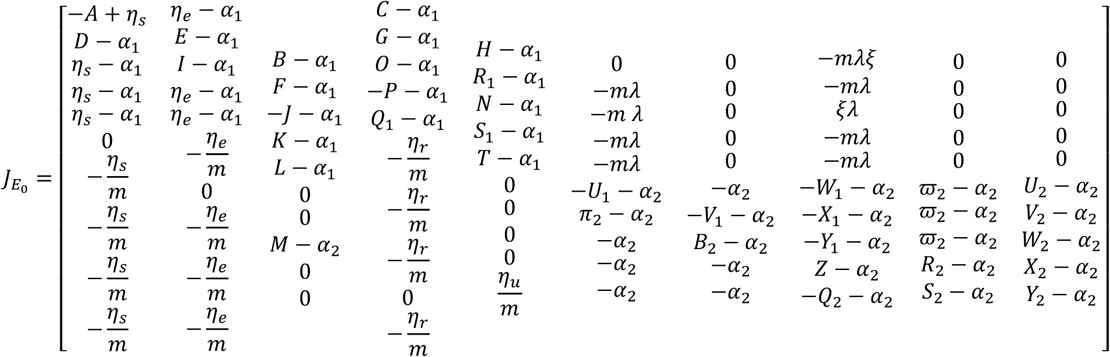

If the Jacobian matrix is evaluated at the disease-free equilibrium state, then the required criteria for stable equilibrium (according to Silver (1999), Theorem 1 and Corollary 3.1) are that the Determinant of the Jacobian 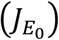 is positive and the Trace of the Jacobian 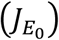 is negative. From the determinant of the Jacobian matrix above given in equation (4.11) we have that

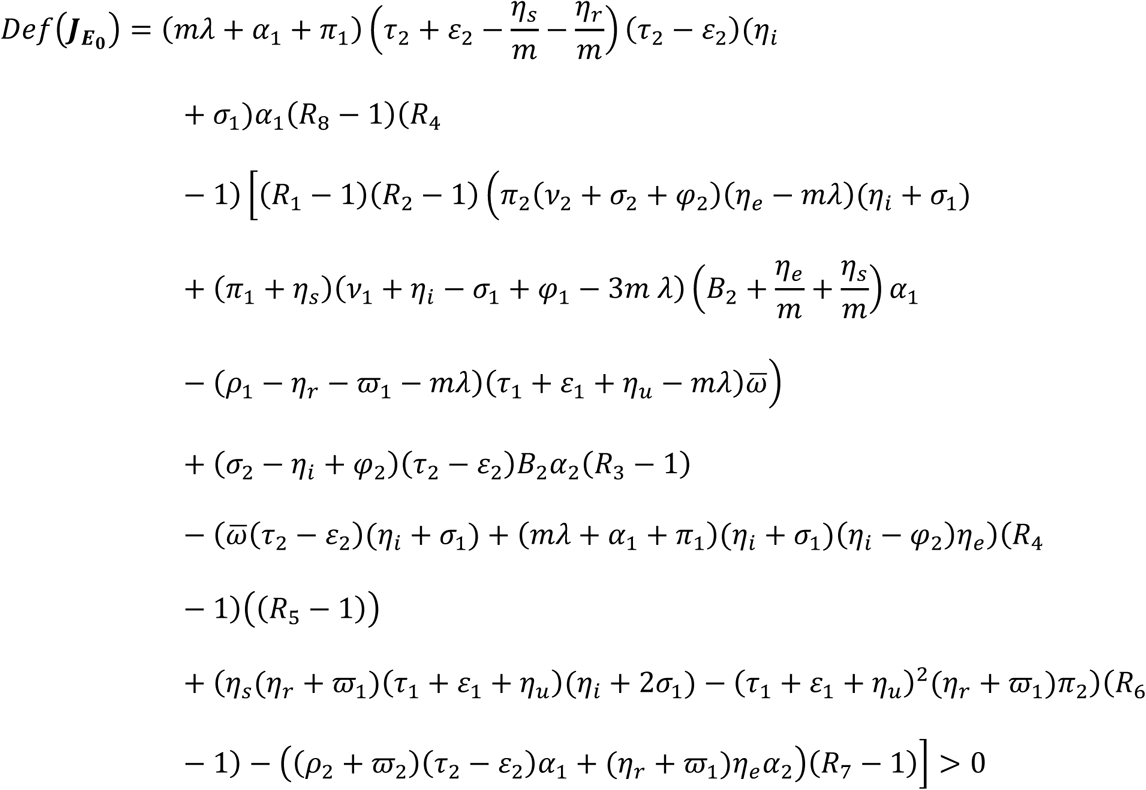

It should be noted however that the following conditions holds: *R*1 < 1, *R*2 < 1, *R*3 < 1, *R*4 < 1, *R*_5_ < 1, *R*_6_ < 1, *R*_7_ < 1 and *R*_8_ < 1, then *Def* 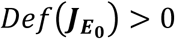, where *R*_1_, *R*_2_, *R*_3_, *R*_4_, *R*_5_, *R*_6_, *R*_7_ and *R*_8_ are defined by equation (4.16).

Similarly from the Trace of the Jacobian matrix 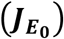 given in equation (4.11) we have that

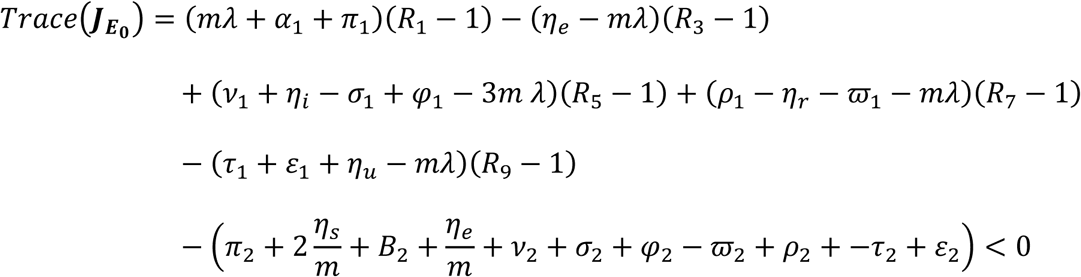

It should be noted that when the following conditions holds *R*_1_ < 1, *R*_3_ < 1, *R*_5_ < 1, *R*_7_ < 1 and *R*_9_ < 1, then *Trace* 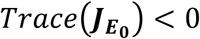 < 0, where *R*_1_, *R*_3_, *R*_5_, *R*_7_ and *R*_9_ are defined in equation (4.16) and (4.18).

Since *Def* 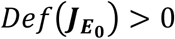 and *Trace* 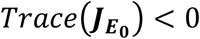 under the prescribed threshold conditions, then the disease free equilibrium **(*E***_**0**_) is locally asymptotically stable.

## 5. CONCLUSION

The investigation of the deterministic endemic SEIRUS model is a novel method in the scientific and empirical approach in controlling the spread of diseases in an endemic situation. This paper focused squarely on determining the asymptotical stability of the new model with the model parameters. We established that a disease-free equilibrium state exists and is locally asymptotically stable when the basic reproduction number 0 ≤ *R*_0_ < 1 and the following threshold conditions (0 ≤ *R*_1_ < 1, 0 ≤ *R*_2_ < 1, 0 ≤ *R*_3_ < 1, 0 ≤ *R*_4_ < 1 and 0 ≤ *R*_5_ < 1, 0 ≤ *R*_6_ < 1, 0 ≤ *R*_7_ < 1, 0 ≤ *R*_8_ < 1) are satisfied. Furthermore, numerical simulations were carried to complement the analytical results in investigating the effect treatment rate and the net transmission rate (that is the probability of secondary transmission) on recovery for both juvenile and adult sub-population in an age-structured population. Results from the model analysis shows that the proportion of infected juvenile and adult sub-population in the presence of HAART drastically reduce to a zero (*R*_0_ = 0) as compared to the infected age-structured population when treatment rate is low and the net transmission rate is near zero. It further shows that the rate of transmission in the adult sub-population from infected mothers to their babies through vertical transmission, contributes highly on the proportion of infections in the newborn babies. Result in this study further shows that high treatment rate with HAART should not be done in isolation, but rather combined with behavioral change to reduce the prevalence of the infection.

## Data Availability

The data referred to in this are available in the manuscript as well as in WPR (2019) and UNAID (2017). Other data are available in CDC (2018). Meanwhile, some of the data are from author's computation using the model parameters defined for the study.

